# The inactivated herpes zoster vaccine HZ/su induces a varicella zoster virus specific cellular and humoral immune response in dialysis patients

**DOI:** 10.1101/2024.05.05.24306698

**Authors:** Franziska Hielscher, Tina Schmidt, Martin Enders, Sarah Leyking, Markus Gerhart, Kai van Bentum, Janine Mihm, David Schub, Urban Sester, Martina Sester

## Abstract

To evaluate the immunogenicity of the inactivated herpes zoster vaccine HZ/su in patients at increased risk for VZV-reactivation, we analyzed the quantity and quality of the vaccine-induced cellular and humoral immunity in dialysis patients with uremic immunodeficiency.

In this observational study, 29 patients and 39 immunocompetent controls underwent standard dual-dose vaccination. Blood samples were analyzed before and two weeks after each vaccination, and after one year. Specific T-cells were characterized after stimulation with VZV-gE peptides based on induction of cytokines and CTLA-4-expression using flow-cytometry. Antibodies were analyzed using ELISA.

Both groups showed an increase in VZV-gE specific CD4 T-cell levels over time (p<0.0001), although median levels reached after second vaccination were lower in patients (0.17% (IQR 0.21%)) than in controls (0.24% (IQR 0.3%), p=0.042). VZV-gE specific CD8 T-cells were only poorly induced. CTLA-4 expression on VZV-gE specific CD4 T-cells was strongest after second dose with no differences between the groups (p=0.45). Multifunctional cells co-expressing IFNɣ, IL-2, and TNF were higher in patients after first vaccination (p=0.028). Median VZV-specific IgG-levels reached a maximum after second vaccination with significantly lower levels in patients (10796 (IQR 12482) IU/l) than in controls (16899 (IQR 14019) IU/l, p=0.009). Despite similar CD4 T-cell levels after one year (p=0.415), antibody levels remained significantly lower in patients (p=0.0008).

The VZV-gE vaccine induced specific antibodies and CD4 T-cells in both patients and controls, whereas CD8 T-cells were only poorly induced. Quantitative and qualitative differences in immunity in patients may indicate reduced duration of protection which may necessitate booster vaccinations.

**Graphical abstract:** 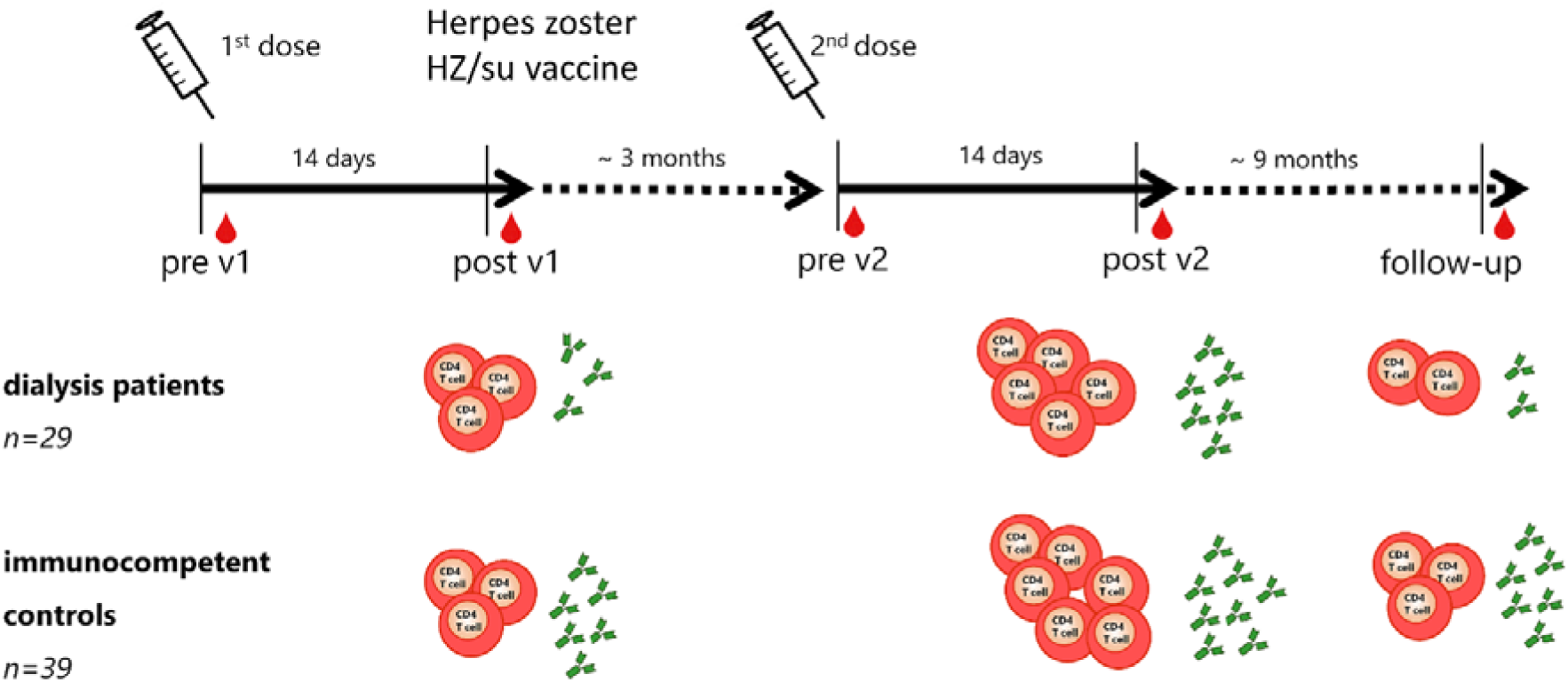

**Lay Summary:** Little is known about the immunogenicity of the inactivated HZ/su in dialysis patients who are at increased risk for VZV reactivation. We therefore analyzed and characterized the cellular and humoral immune response induced by HZ/su in dialysis patients compared to healthy individuals. HZ/su induces VZV-specific CD4 T-cells and antibodies in both controls and dialysis patients, whereas VZV-specific CD8 T-cells were only poorly induced. VZV-specific CD4 T-cells were multifunctional and showed a dynamic increase with a maximum after the second vaccination. However, median T-cell levels were lower in patients. Also VZV-specific IgG antibodies showed a dynamic increase in both groups, although after second vaccination and one year after vaccination antibody levels of patients were lower compared to controls. Future studies should address whether differences in quantity and quality of vaccine-induced VZV-specific T-cells and lower antibody levels in patients may indicate a reduced protective effect, which may necessitate booster vaccinations.

## Introduction

Varicella zoster virus (VZV), a member of the alphaherpesvirus family, causes chickenpox during primary infection in childhood, and then remains latent in dorsal root ganglia or cranial nerves. VZV may reactivate from latency causing herpes zoster (HZ)^1^, which typically manifests as a painful, dermatomal vesicular rash and may lead to serious complications such as postherpetic neuralgia (PHN)^2, 3^. The probability of reactivation depends on the individual’s immunity, whereby reduced VZV-specific T-cell levels are considered to be the main determinant. A decline in VZV-specific T-cell immunity with increasing age is associated with an increased risk of VZV reactivation among the elderly^4, 5^. The incidence of HZ is about 4-4.5 per 1,000 person-years and increases to more than 11 per 1,000 person-years in people aged 80 years or older^6^. Another factor that favors VZV reactivation and increases the risk of herpes zoster is an impaired immune system^7, 8^. Dialysis patients with uremic immune dysfunction also belong to this risk group, which has a higher probability of VZV reactivation than immunocompetent individuals^9, 10^. These patients often exhibit a disturbed interaction between APCs and T-cells and an increased production of pro-inflammatory cytokines^11^. This is associated with more frequent and severe infections^12^ as well as an inadequate vaccination response, e.g. against hepatitis B, tetanus^13^, diphtheria^14^ and influenza^15, 16^.

In 2018, a recombinant glycoprotein E (gE) subunit herpes zoster vaccine (HZ/su) was approved in Germany^17, 18^. Unlike the previously available live-attenuated vaccine Zostavax, HZ/su is also suitable for immunocompromised people for whom the live-attenuated vaccine is normally contraindicated^19^. HZ/su consists of VZV-gE and AS01_B_ as adjuvant system, which contains *Quillaja saponaria* Molina, fraction 21 (QS-21) and 3-O-desacyl-4′-monophosphoryl lipid A (MPL) from *Salmonella Minnesota*^20^. It has already been shown to be effective for healthy individuals^21, 22^ or patients after autologous stem cell transplantation^23^. Nevertheless, we hypothesized that immunogenicity may be impaired in immunocompromised patients. First immunogenicity data exist in patients after solid organ transplantation^24^. However, a direct comparison with controls was not performed, and knowledge on the vaccine-induced immune response and its stability in dialysis patients is limited. In this observational study, we therefore aimed at characterizing the reactogenicity and immunogenicity of HZ/su in dialysis patients compared to healthy individuals. In addition to the quantitative determination of VZV-specific T-cells and antibodies, a qualitative investigation of proliferative capacity, cytokine expression of vaccine-induced T-cells and neutralizing effect of the specific antibodies was performed.

## Methods

### Recruitment of the study population

Dialysis patients and immunocompetent age-matched controls without history of herpes zoster vaccination receiving two standard dosages of the HZ/su vaccine based on standard recommendations were enrolled in an observational study from 06/2019 to 12/2021. Patients had been on dialysis for at least 6 months. Whole blood samples were collected before the first and second HZ/su vaccination, two weeks after each vaccine dose as well as 12 months after the first (i.e. 9 months after the second vaccination; supplementary figure S1). Blood samples were drawn before the dialysis sessions. Study participants completed a questionnaire for self-reporting of local and systemic adverse events occurring within the first week after each vaccination. We anticipated a sample size of approximately 30 individuals per group based on previous immunogenicity studies^15, 25^. The study was approved by the ethics committee of the Ärztekammer des Saarlandes (reference 27/19), and all individuals gave written informed consent.

### Quantification of lymphocyte subpopulations

To quantitate lymphocyte subpopulations, 100µl heparinized whole blood was washed once with RPMI, stained for characteristic phenotypic markers and analyzed using flow cytometry as described before^26^. Details on the gating strategy and on the antibodies are given in the supplement.

### Quantification and characterization of VZV-specific T-cells

The analysis of antigen-specific T-cells was carried out as previously described for other VZV antigens^27, 28^. In brief, 2µg/ml overlapping VZV gE peptides (Swiss-Prot ID: P09259, JPT, Berlin, Germany) and 2.5 μg/ml *Staphylococcus aureus* enterotoxin B (SEB; Sigma-Aldrich, St. Louis, MO, USA) were used for stimulation. After 6 hours, VZV-specific CD4 or CD8 T-cells were identified using flow-cytometry as activated CD69-positive T-cells producing IFNγ, and further characterized for expression of cell surface markers and additional cytokines. Details on the stimulatory procedure and on the antibodies are given in the supplement.

### Proliferation activity of VZV-specific T-cells

VZV-specific proliferation was analyzed using carboxyfluoresceindiacetate-succinimidylester (CFDA-SE) assay, as described before^29^ and in the supplement.

### Analysis of VZV-specific IgG antibodies and neutralization activity

VZV-specific antibodies were quantified using a commercial anti-IgG enzyme-linked immunosorbent assay according to the manufactureŕs instructions (Euroimmun AG, Lübeck, Germany). The functionality of VZV-specific antibodies was characterized by a neutralization test based on VZV infection (Clade 3, strain Nr. 13, Original-Nr: 1219/07) of embryonic lung fibroblasts. Details on the experimental procedure are given in the supplement.

### Statistical analysis

The Mann-Whitney test was used for analysis of nonparametric data between two groups. Parametric analyses were performed using the unpaired t-test. Longitudinal analyses of paired samples were performed using the Friedman test with Dunńs post-test or the Wilcoxon test. Fisher’s exact test and Chi-square test was used to analyze differences in categorical parameters. Correlations were calculated using the nonparametric Spearman test. Statistical analysis was performed using GraphPad-Prism-V9.2.0 (GraphPad, San Diego, CA). A p-value <0.05 was considered statistically significant.

## Results

### Study population

A total of 29 dialysis patients and 39 healthy controls were recruited, who received the dual dose inactivated HZ/su vaccine. Demographic characteristics are shown in table 1, with underlying diseases of dialysis patients listed in supplementary table S1. Most subjects reported a history of chickenpox (controls 91.2%; patients 76.2%), and 24.0% of patients and 18.4% of controls had already suffered from herpes zoster. Patients and controls differed in their leukocyte subpopulations (table 1). Monocyte and granulocyte counts were significantly higher in dialysis patients (p=0.006 and p=0.035, respectively), whereas lymphocyte counts were significantly lower (p<0.0001). While CD8 T-cell counts did not differ in both groups, dialysis patients had significantly lower CD4 T-cell counts (p=0.031). In addition, numbers of CD19-positive B-cells (p<0.0001) and plasmablasts (p=0.004), identified as CD38-positive cells among IgD^−^CD27^+^ CD19-positive switched-memory B-cells, were significantly lower in dialysis patients.

**Table 1:**
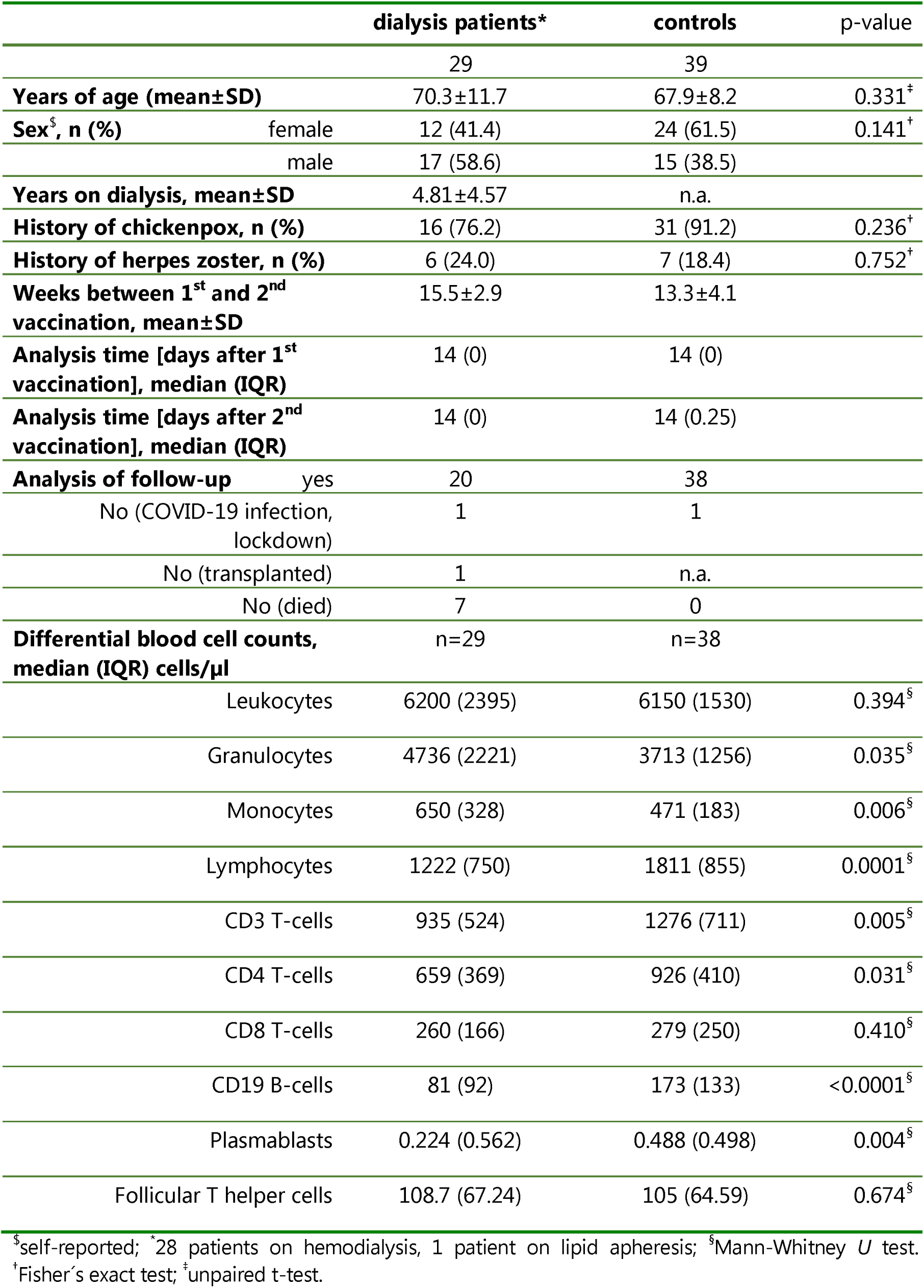
Demographic and basic characteristics of the study populations.

### HZ/su vaccine mainly induces VZV-specific CD4 T-cells with only low levels of CD8 T-cells

The induction of the VZV-specific immune response by the HZ/su vaccine was analyzed in patients and controls immediately before each vaccination (pre v1 and pre v2), 2 weeks after (post v1 and post v2) and 12 months after the first vaccination (follow-up, supplementary figure S1). VZV-specific T-cells were identified using flow cytometry based on the induction of the activation marker CD69 and the cytokine IFNγ after specific stimulation with overlapping VZV gE peptides. Diluent (DMSO) and *Staphylococcus aureus* Enterotoxin B (SEB) was used as negative and positive control stimuli, respectively. Representative contour plots of blood samples of a male hemodialysis patient in his 50s after peptide stimulation is shown in figure 1a. A significant increase in vaccine-specific CD4 T-cells was observed two weeks after the first and the second vaccination in both patients and controls, with the maximum peak after the second vaccination (figure 1b). At follow-up, VZV-specific CD4 T-cell levels decreased again, but remained higher than CD4 T-cell frequencies prior to vaccination (p<0.0001). In contrast, neither patients nor controls showed a significant increase in VZV-specific CD8 T-cell levels after the two vaccinations. In some cases, VZV-specific CD8 T-cell levels were high even before vaccination, and remained stable over time. Despite similar dynamics in VZV-specific CD4 T-cells in both groups, patients reached significantly lower VZV-specific CD4 T-cell levels (0.17% (IQR 0.21%)) two weeks after the second vaccination compared to healthy controls (0.24% (IQR 0.3%) p=0.042, figure 1c). Likewise, the median increase in the percentage of specific CD4 T-cells from baseline to two weeks after the second vaccination was lower in dialysis patients (7.7-fold) than in controls (23.3-fold; p=0.010). Finally, at one year follow-up, the increase in patients was significantly lower than in controls (2.9-fold versus 6.6-fold; p=0.048, figure 1d). Overall, dynamics in T-cell levels were vaccine-specific, as the percentage of SEB-reactive CD4 and CD8 T-cells were largely similar in the two groups and remained stable over time (figure 1b).

**Figure 1:**
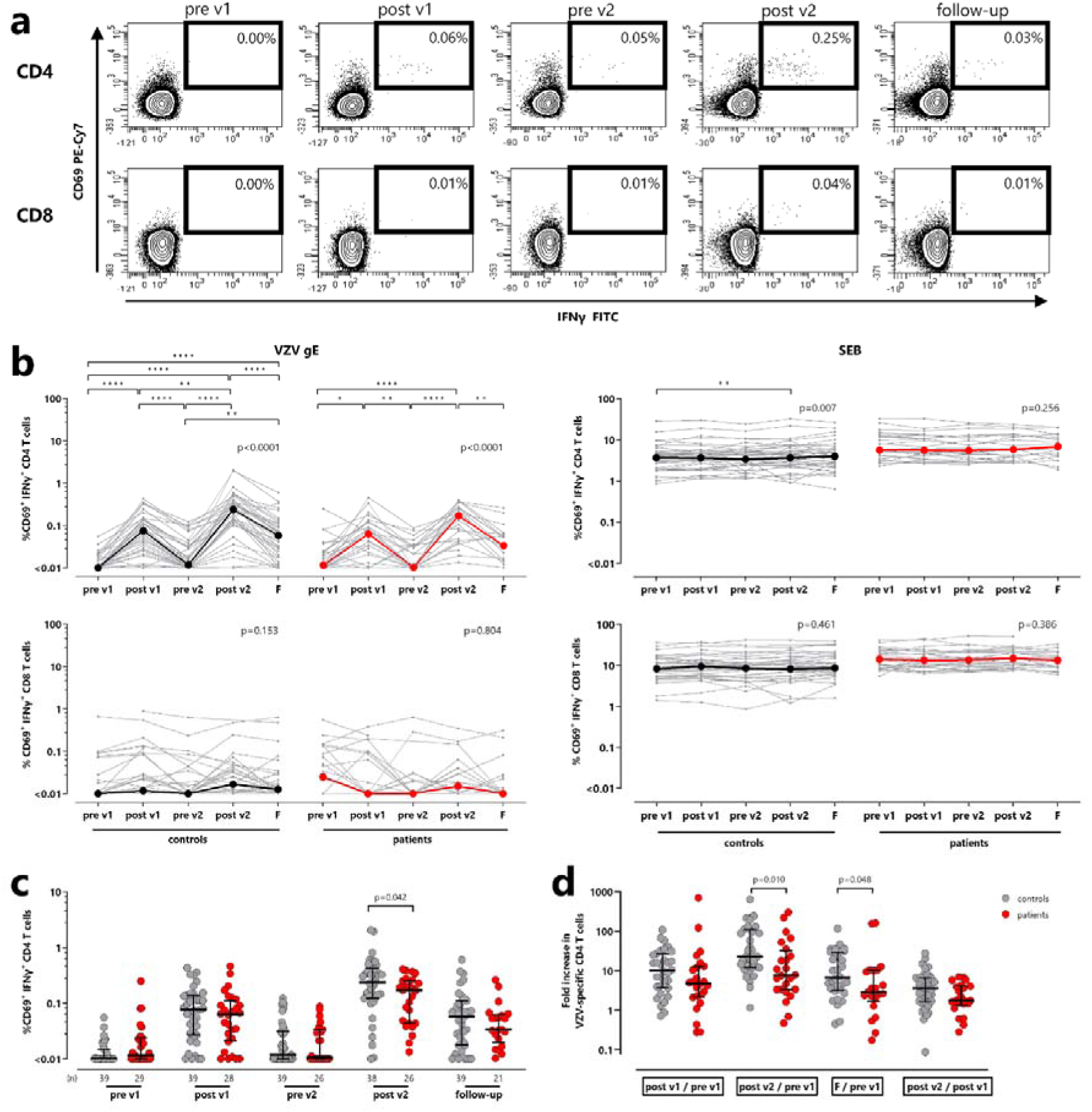
VZV-specific CD4 T-cell levels increase after HZ/su vaccination. **(a)** Representative contour plots of specific CD4 and CD8 T-cells of a male dialysis patient in his 50s before the first and the second vaccinations (pre v1; pre v2), two weeks after the first and the second vaccinations (post v1; post v2) as well as 9 months after the second vaccination (follow-up) determined after stimulation of whole blood with overlapping peptides of VZV gE. Numbers indicate the percentages of reactive CD4 and CD8 T-cells defined by co-expression of the activation marker CD69 and the cytokine IFNγ. **(b)** Percentages of VZV-specific CD4 (upper panels) and CD8 T-cells (lower panels) after subtraction of the corresponding negative control (left) and SEB-reactive CD4 and CD8 T-cells (right) over time. Bold lines represent median values. Friedman test with Dunńs post test was performed for statistical analysis. **(c)** Comparison of samples from healthy controls (red) and dialysis patients (grey) at each time point. Bars represent median values with interquartile ranges. **(d)** For each individual the fold increase in VZV-gE specific CD4 T-cell levels was calculated after the first and second vaccination and at follow-up compared with baseline (pre v1) and between the first and the second vaccination (post v1/pre v1, post v2/pre v1, follow-up/ pre v1 and post v2/post v1). Statistical analysis in C and D was performed using Mann-Whitney test. F, follow-up; IFN, interferon; VZV, *Varicella zoster virus*; SEB, *Staphylococcus aureus* enterotoxin B.

### Vaccine-induced changes in CTLA-4 and cytokine expression in VZV-specific CD4 T-cells

VZV-specific CD4 T-cells were analyzed for expression of CTLA-4 after both vaccinations as marker for recent antigen encounter. Contour plots of the CTLA-4 expression of VZV-specific and SEB-reactive CD4 T-cells from a male dialysis patient in his 50s two weeks after the second vaccination are shown in figure 2a. CTLA-4 expression levels of VZV-specific CD4 T-cells were higher than of SEB-reactive CD4 T-cells (figure 2b). Moreover, CTLA-4 expression of VZV-specific CD4 T-cells was numerically higher after the second vaccination than after the first, although this difference only reached statistical significance in controls (p=0.013 vs. p=0.064 in patients). CTLA-4 expression on follow-up decreased in both groups (figure 2c). When comparing patients and controls, no difference in CTLA-4 expression was found neither after the first nor the second vaccination (figure 2c).

**Figure 2:**
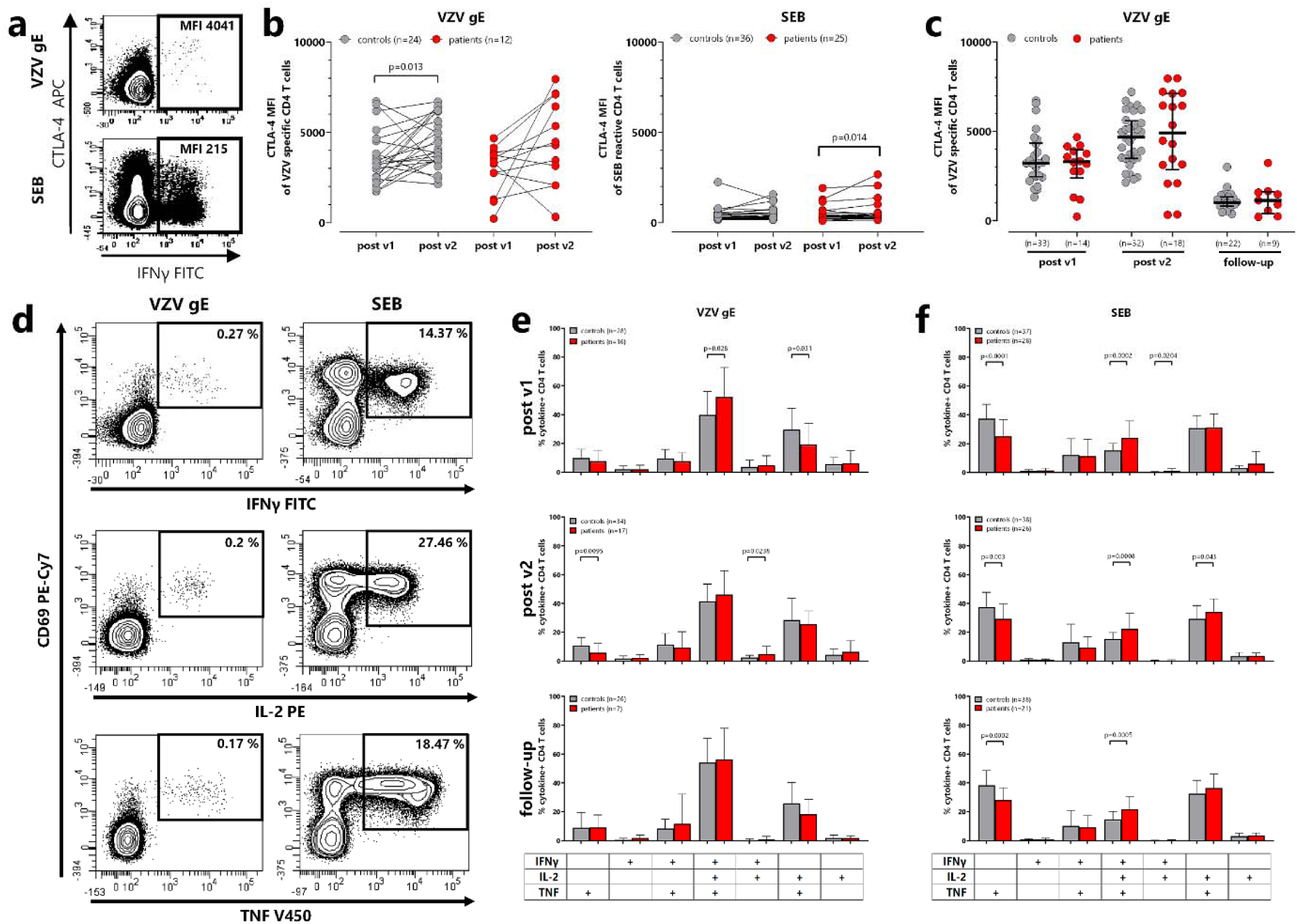
CTLA-4 expression and cytokine profile of VZV-specific CD4 T-cells after Hz/su vaccination. **(a)** Representative contour plots of median fluorescence intensity (MFI) of CTLA-4 expressing VZV-specific CD4 T-cells of a male dialysis patient in his 50s. **(b)** Comparison of CTLA-4 MFI of VZV-specific and SEB-reactive cells after the first and after the second vaccination in controls (grey, p=0.013) and dialysis patients (red, p=0.064). All samples were measured. To allow for robust statistical analysis, only paired samples with at least 20 CD69+IFNγ+ CD4 T-cells were included. Differences between the time points in each group were calculated using the Wilcoxon test. **(c)** Comparison of CTLA-4 MFI on VZV-specific CD4 T-cells after both vaccinations, and on follow-up between controls and dialysis patients. Statistical analysis was performed using the Mann-Whitney test. **(d)** Examples of contour plots of VZV-specific or SEB-reactive CD4 T-cells expressing cytokines interferon gamma (IFNγ), tumor necrosis factor (TNF) and interleukin 2 (IL-2) after stimulation of a whole blood sample from a male dialysis patient in his 50s. Cytokine expressing CD4 T-cells were subclassified into 7 subpopulations according to single or combined expression of IFNγ, TNF and IL-2. Blood samples from all individuals were analyzed. To ensure robust statistics, only samples with at least 30 cytokine-expressing CD4 T-cells after subtraction of the corresponding negative control stimulation were considered. Comparison of cytokine profiles of **(e)** VZV-specific and **(f)** SEB-reactive CD4 T-cells in dialysis patients and controls post v1, postv2, and on follow-up time points. Bars represent means and standard deviations. Statistical analysis was performed using unpaired t-test. The final sample size is indicated in each panel. CTLA-4, cytotoxic T-lymphocyte antigen 4; IFN, Interferon; IL, Interleukin; VZV, Varicella zoster virus; SEB, *Staphylococcus aureus* Enterotoxin B; TNF, tumor necrosis factor.

In addition, the expression profiles of the cytokines IFNγ, IL-2 and TNF were analyzed. Representative contour plots of CD69-positive VZV-specific and SEB-reactive CD4 T-cells producing the individual cytokines are shown in figure 2d, with quantitative analyses for all individuals displayed in supplementary figure S2. As with IFNγ-producing cells (figure 1c), the most pronounced differences between patients and controls were found after the second vaccination. Subdivision of cytokine-producing cells by boolean gating resulted in a total of seven subpopulations defined by expression of three cytokines, two cytokines or one cytokine only. VZV-specific CD4 T-cells are characterized by multifunctionality with the majority of cells simultaneously expressing all three cytokines (figure 2e), which contrasts with SEB-reactive CD4 T-cells, which predominantly express only TNF or TNF in combination with IL-2 (figure 2f). After the first vaccination, the percentage of triple positive VZV-specific CD4 T-cells was significantly higher in patients than in controls (p=0.028), while the percentage of IL-2/TNF-expressing cells was concomitantly lower (p=0.031). Overall, cytokine expression patterns remained similar after the second vaccination and on follow-up.

### Differentiation status of VZV-specific CD4 T-cells after vaccination

To analyze the differentiation status of VZV-specific T-cells, CD69^+^ IFNγ^+^ CD4 T-cells were classified into naive, central memory (CM), effector memory (EM), and terminally differentiated effector memory (TEMRA) cells based on expression of CD45RO and CD27 after stimulation with overlapping VZV gE peptides (figure 3a). After the first vaccination, central memory T-cells accounted for the largest proportion of VZV-specific CD4 T-cells in both dialysis patients (80.8% (IQR 20.3%)) and controls (76.0% (IQR 20.3%)), followed by effector memory T-cells (patients: 18.2% (IQR 22.8%); controls 14.5% (IQR 19.8%), whereas the proportion of TEMRA or naive CD4 T-cells was very low. A similar distribution was also observed two weeks after the second vaccination and on follow-up, with some differences between patients and controls after the second vaccination. While patients had a higher percentage of central memory T-cells (p=0.003), the percentage of effector memory T-cells was concomitantly lower than in controls (p=0.004, figure 3b). Although the majority of SEB-reactive T-cells also had a central memory phenotype, the pattern was distinct from VZV-specific CD4 T-cells with no differences between patients and controls (figure 3c). Moreover, the distribution of VZV-specific subpopulations was distinct from bulk CD4 and CD8 T-cells (supplementary figure S3). Among bulk CD4 T-cells, we also characterized Tfh-cells in circulation which were predominantly of a central memory phenotype, but their levels and phenotype did not show any pronounced vaccine-related changes over time (supplementary figure S4).

**Figure 3:**
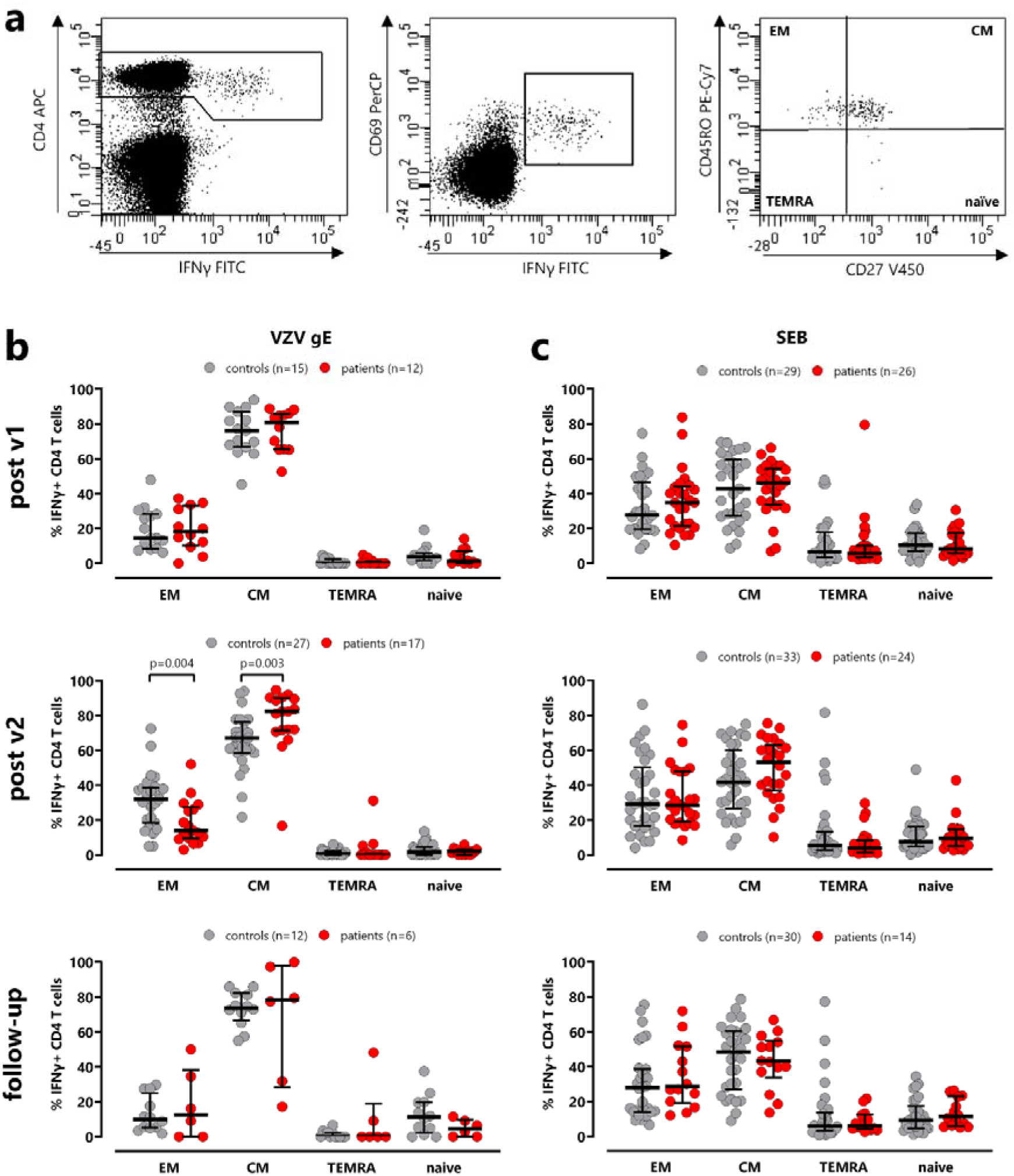
Differences in VZV-specific CD4 differentiation between dialysis patients and controls. **(a)** Representative contour plots of the differentiation status of VZV-specific CD4 T-cells, identified by CD4 T-cells expressing CD69 and IFNγ and further classified into naïve, central memory (CM), effector memory (EM), and terminally differentiated effector memory (TEMRA) cells based on expression of CD45RO and CD27. **(b)** T-cell populations among VZV-specific or **(c)** SEB-reactive CD4 T-cells were compared between controls (gray) and dialysis patients (red) after the first and the second vaccinations and on follow-up. All samples were measured, but the final analysis was restricted to samples with at least 20 CD69+IFNγ+ CD4 T-cells to ensure robust statistical analysis. The final sample size is indicated in each panel. Statistical analysis was performed using Mann-Whitney test. Bars represent medians with interquartile ranges.

### HZ/su induced proliferation activity in VZV-specific T-cells

Apart from cytokine-producing cells after short-term stimulation, the proliferation capacity of VZV-specific CD4 and CD8 T-cells after antigen-specific stimulation was examined using a CFDA-SE assay. Representative contour plots of proliferating CD4 and CD8 T-cells after staining with CFDA-SE and stimulation with overlapping VZV gE peptides, DMSO diluent as negative control and SEB as positive control for 7 days are shown in figure 4a. Two weeks after each vaccination, an increase in the percentage of proliferating VZV-specific CD4 T-cells was observed in both dialysis patients and controls. In addition, the proliferative capacity of VZV-specific CD4 T-cells remained higher in the follow-up samples than in samples before the first vaccination (figure 4b). Interestingly, VZV-specific CD8 T-cell proliferation was also induced, although the proliferative capacity was lower than that of CD4 T-cells (figure 4b). As expected, proliferation of SEB-reactive CD4 and CD8 T-cells was stable over time (figure 4c). The percentage of proliferating T-cells as well as the increase in VZV-specific T-cells were largely similar in both groups, except that the proliferative capacity of VZV-specific CD8 T-cells on follow-up was slightly lower in patients (p=0.047, figure 4d and e).

**Figure 4:**
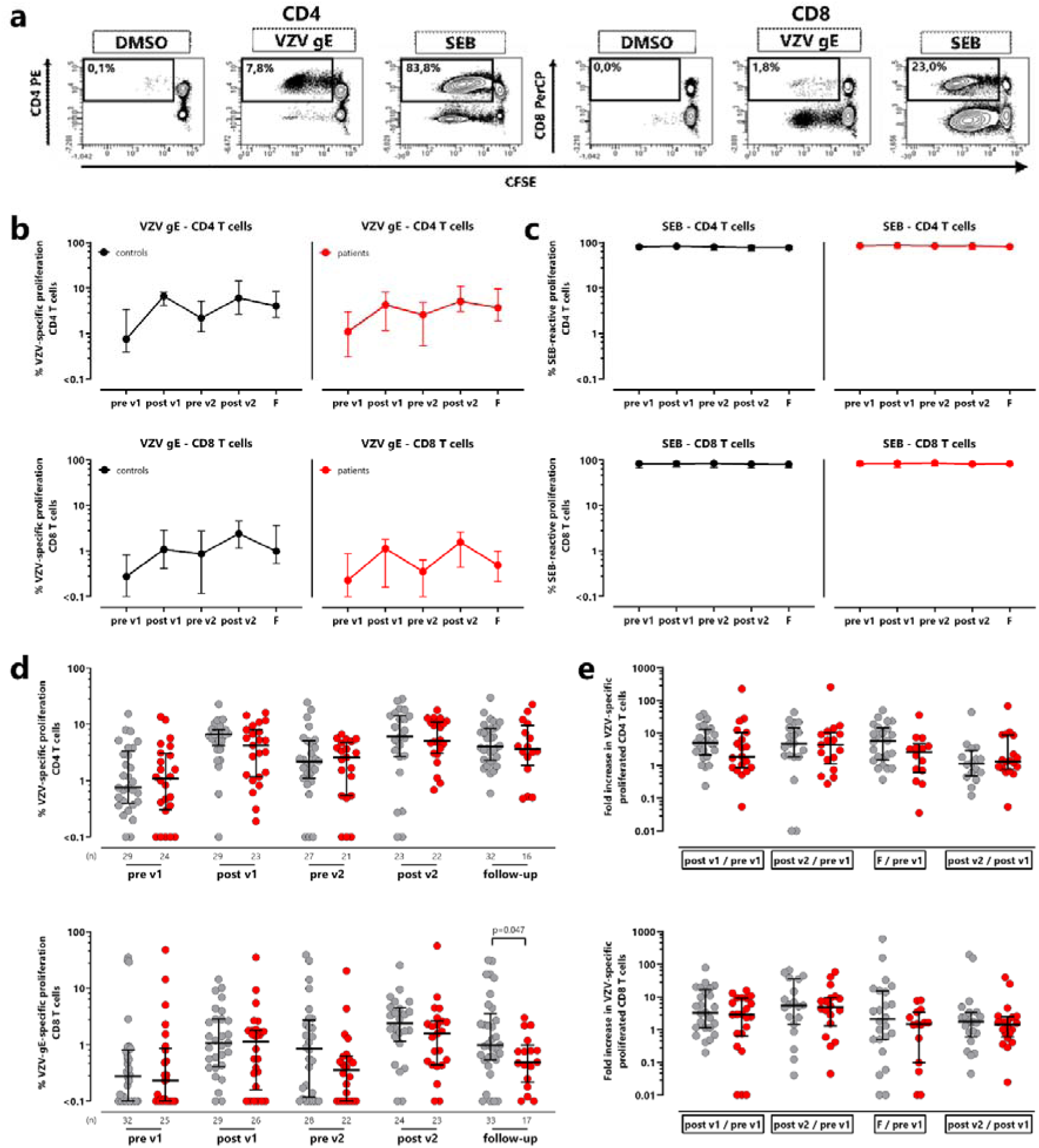
Induction of VZV-specific T-cell proliferation in dialysis patients after vaccination with HZ/su. **(a)** Contour plots of proliferated CD4 and CD8 T-cells after staining with CFDA-SE and stimulation with overlapping VZV gE peptides, SEB (positive control) or DMSO (negative control) for 7 days. **(b)** Median percentages and IQR of proliferating VZV-specific CD4 (upper panel) or CD8 T-cells (lower panel) or **(c)** SEB-reactive CD4 or CD8 T-cells before and after vaccinations. VZV-specific CD4 and CD8 T-cells are shown after subtraction of the corresponding negative control over time in dialysis patients (red) in comparison to healthy controls (gray). **(d)** The percentage of proliferated VZV-specific CD4 and CD8 T-cells from patients and controls are compared at each time point. **(e)** For each individual the fold increase of proliferated VZV-specific CD4 and CD8 T-cell level was calculated after the first and second vaccination and at follow-up compared with baseline and between the first and second vaccination (post v1/pre v1, post v2/pre v1, follow-up/ pre v1 and post v2/post v1). Bars refer to medians with interquartile ranges and Mann-Whitney test was used for statistical analysis.

### Differences in B-cell subpopulations of dialysis patients compared to healthy individuals

We next analyzed B-cells and their subpopulations which were divided into naive, non-switched and switched memory B-cells based on expression of IgD and CD27. Moreover, plasmablasts were quantified as CD38-positive cells among switched memory B-cells (figure 5a). In both dialysis patients and controls, the percentage of plasmablasts among B-cells was low and remained stable over time with no vaccine-associated dynamics (controls: p=0.901; patients: p=0.063, figure 5b). When analyzing the distribution of the subpopulations over time, most B-cells showed a naive phenotype at all time points (figure 5c). Interestingly, the proportion of naive B-cells was higher in dialysis patients than in controls, which reached statistical significance two weeks after the second vaccination (p=0.045). In contrast, dialysis patients showed a significantly lower percentage of non-switched B-cells compared to controls throughout the observation period (pre v1: p=0.011; post v2: p=0.011; follow-up: p=0.004, figure 5c).

**Figure 5:**
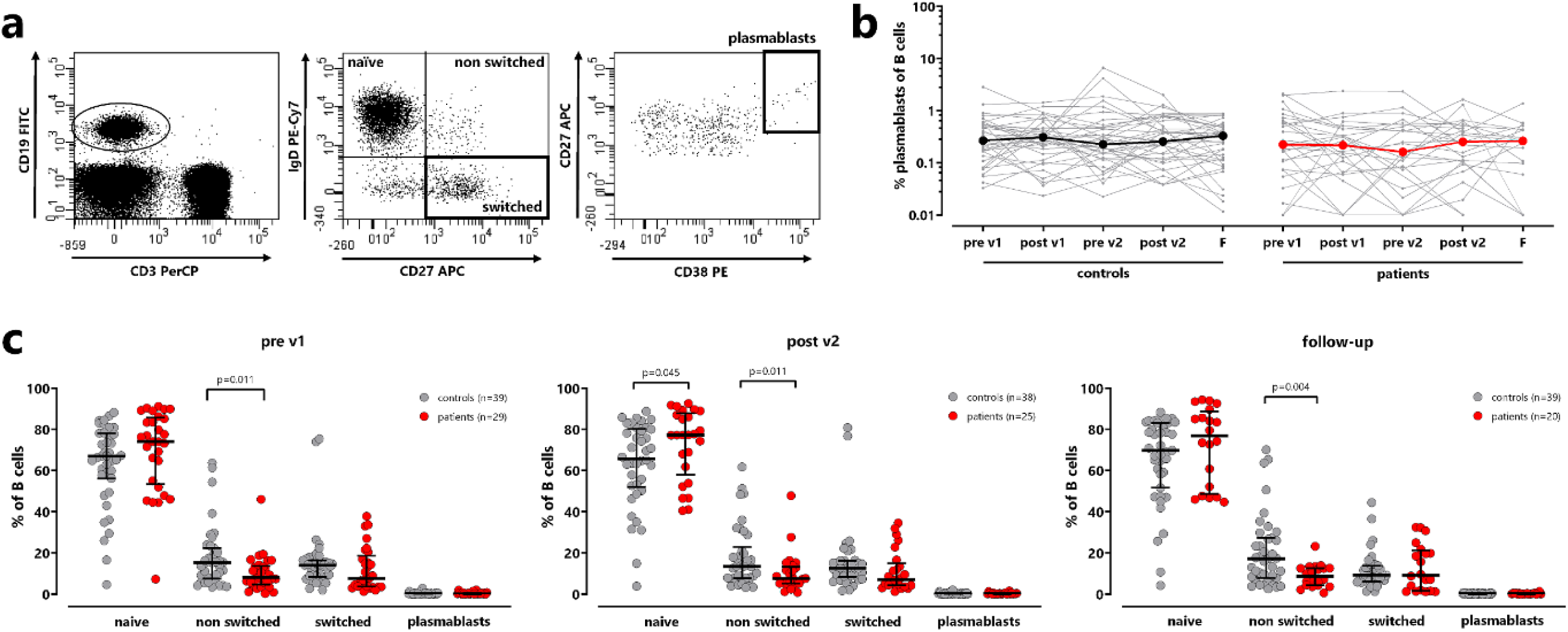
Higher proportion of naïve B cells in dialysis patients. **(a)** Representative dotplots of CD19+ B-cells, classified into naïve, non-switched, and switched memory B-cells based on the surface markers IgD and CD27. Plasmablasts were identified as CD38-positive cells among switched memory B-cells. **(b)** The percentage of plasmablasts was determined over time in controls (left) and patients (right). Bold lines represent median values. **(c)** B-cell subpopulations of controls (grey) and dialysis patients (red) were compared before the first vaccination (pre v1), after the second vaccination (post v2) and on follow-up. Bars represent median values with interquartile ranges. Statistical analysis was performed using Mann-Whitney test.

### VZV-specific humoral immune response is lower in dialysis patients

To investigate the vaccine-induced humoral immune response, VZV-specific IgG levels were determined by ELISA. In total, only 2 dialysis patients and 4 healthy controls were seronegative before vaccination (<80 IU/l), while all other participants had levels above detection limit. As with specific CD4 T-cells, VZV-specific IgG antibodies showed a dynamic increase in both groups (figure 6a). Two weeks after each vaccination, an increase in IgG concentration was observed in dialysis patients (post v1: 9139 (IQR 9582) IU/l; post v2: 10796 (IQR 12482) IU/l) and controls (post v1: 11843 (IQR 12231) IU/l; post v2: 16899 (IQR 14019) IU/l), with peak levels reached after the second vaccination. VZV IgG levels of dialysis patients were significantly lower than those of controls at this time point (p=0.009). Dialysis patients also had significantly lower VZV-specific IgG levels on follow-up (p=0.0008, figure 6b). Consequently, the median increase of VZV-specific IgG from baseline to two weeks after second vaccination was also lower in dialysis patients (5-fold) than in controls (9.1-fold; p=0.004), as was the increase from baseline to follow-up (patients: 1.6-fold; controls: 5.4-fold; p<0.0001), or the increase from the first to the second vaccination (p=0.008, figure 6c).

**Figure 6:**
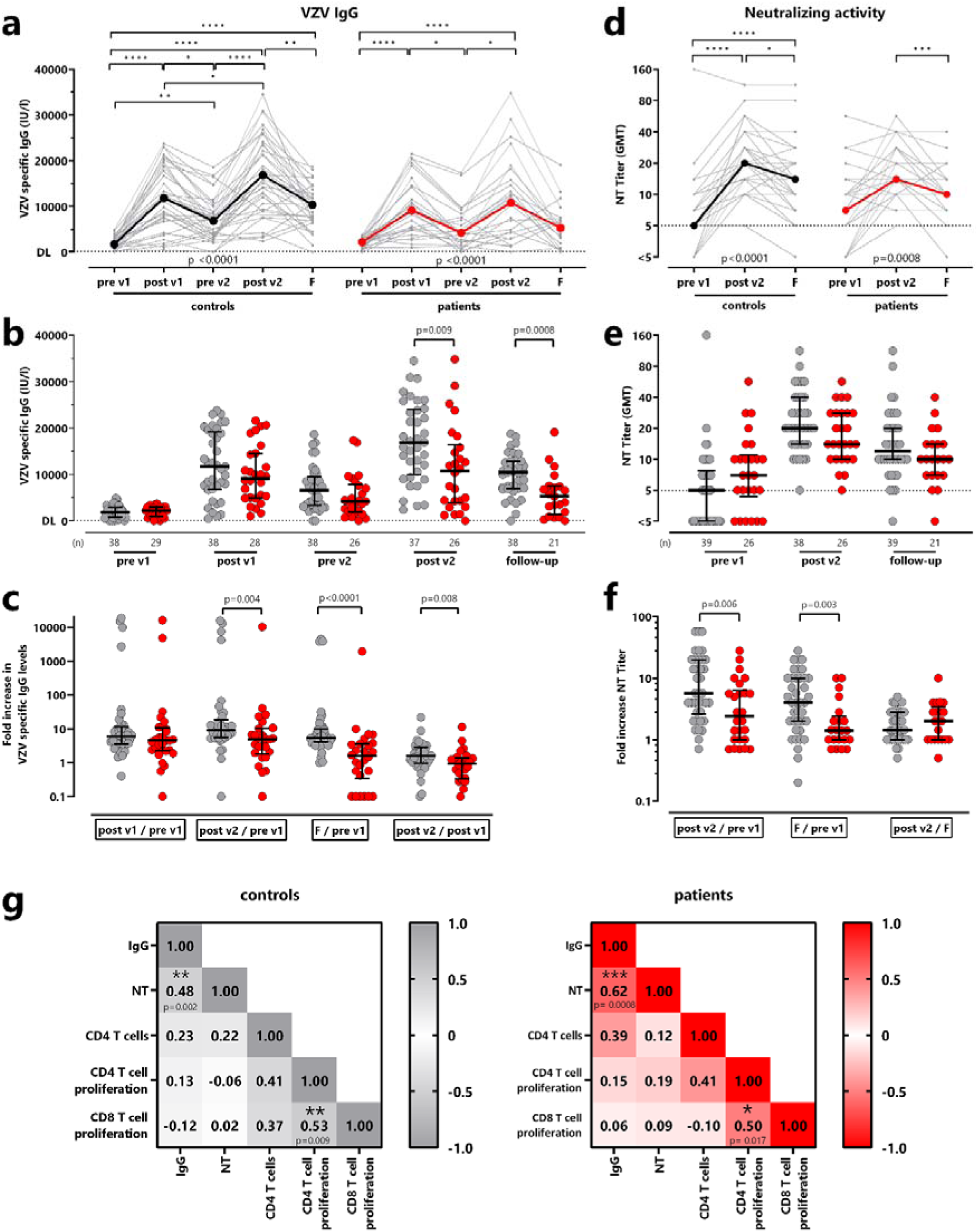
Lower increase in VZV-specific IgG levels and neutralizing antibody titers in dialysis patients. **(a)** VZV IgG levels measured over time in controls (grey) and dialysis patients (red)**. (b)** Comparison of VZV-specific IgG levels between both groups at each time point. **(c)** For each individual the fold increase in VZV-specific IgG levels was calculated after the first and second vaccination and at follow-up compared with baseline and between the first and second vaccination (post v1/pre v1, post v2/pre v1, follow-up/ pre v1 and post v2/post v1). Samples <1 IU/l set as 1 to calculate the fold increase. **(d)** VZV-specific neutralization in dialysis patients (red) and controls (black) is shown after both vaccinations (post v1; post v2) and after one year (follow-up). **(e)** Comparison of neutralizing activity and **(f)** fold increase, calculated after second vaccination and follow-up compared with first vaccination and between second vaccination and follow-up, between both groups. Samples with NT titer <5 set as 1 to calculate the fold increase. Statistical analysis of longitudinal samples was performed using the Friedman test and bold lines represent medians. Statistical analyses on the differences between the patients and controls were performed using the Mann-Whitney test, and bars represent medians with interquartile ranges. **(g)** Correlation matrix between the percentage of vaccine-induced VZV-specific CD4 T-cells (derived from figure 1), IgG levels, neutralizing activity, and the percentage of proliferating VZV-specific CD4 and CD8 T-cells (derived from figure 4) in controls (left) and dialysis patients (right) after second vaccination. Correlation coefficients were calculated according to two-tailed Spearman and displayed using a color code.

Vaccination-induced neutralizing antibody titers were analyzed before the first vaccination, two weeks after the second vaccination and on follow-up and reached their maximum after the second vaccination in both groups. Despite a subsequent decrease, neutralizing activity on follow-up was still higher than before the first vaccination (figure 6d). Although no differences in neutralizing activity was found between the two groups (figure 6e), dialysis patients showed significantly less pronounced increases in neutralizing activity from baseline to two weeks after the second vaccination (p=0.006) and from baseline to follow-up (p=0.003, figure 6f).

A correlation matrix was used to investigate the relationship between the magnitudes of vaccine induced IgG, neutralizing activity, CD4 T-cells and proliferative capacity in both groups after the second vaccination. A significant correlation was observed between VZV-specific IgG levels and neutralizing activity in both controls (r=0.48, p=0.002) and patients (r=0.62, p=0.0008). Moreover, both groups showed a correlation between the proliferative capacity of VZV-specific CD4 and CD8 T-cells (controls: p=0.009; patients: p=0.017, figure 6g).

### Low reactogenicity after HZ/su vaccination in dialysis patients

Vaccine-related adverse events were compared between patients and controls in the first week after both vaccinations based on self-reporting using a questionnaire. Overall, both vaccinations were well tolerated with pain at the injection site followed by redness at the injection site and fatigue being most frequently reported (figure 7a). Compared with healthy controls, dialysis patients reported fewer local and/or systemic adverse events (figure 7b). In general, most controls reported similarly frequent adverse events after the first and the second vaccination. In contrast, adverse events among patients tended to be less frequent after the second vaccination, with the exception of fatigue and headache (figure 7c).

**Figure 7:**
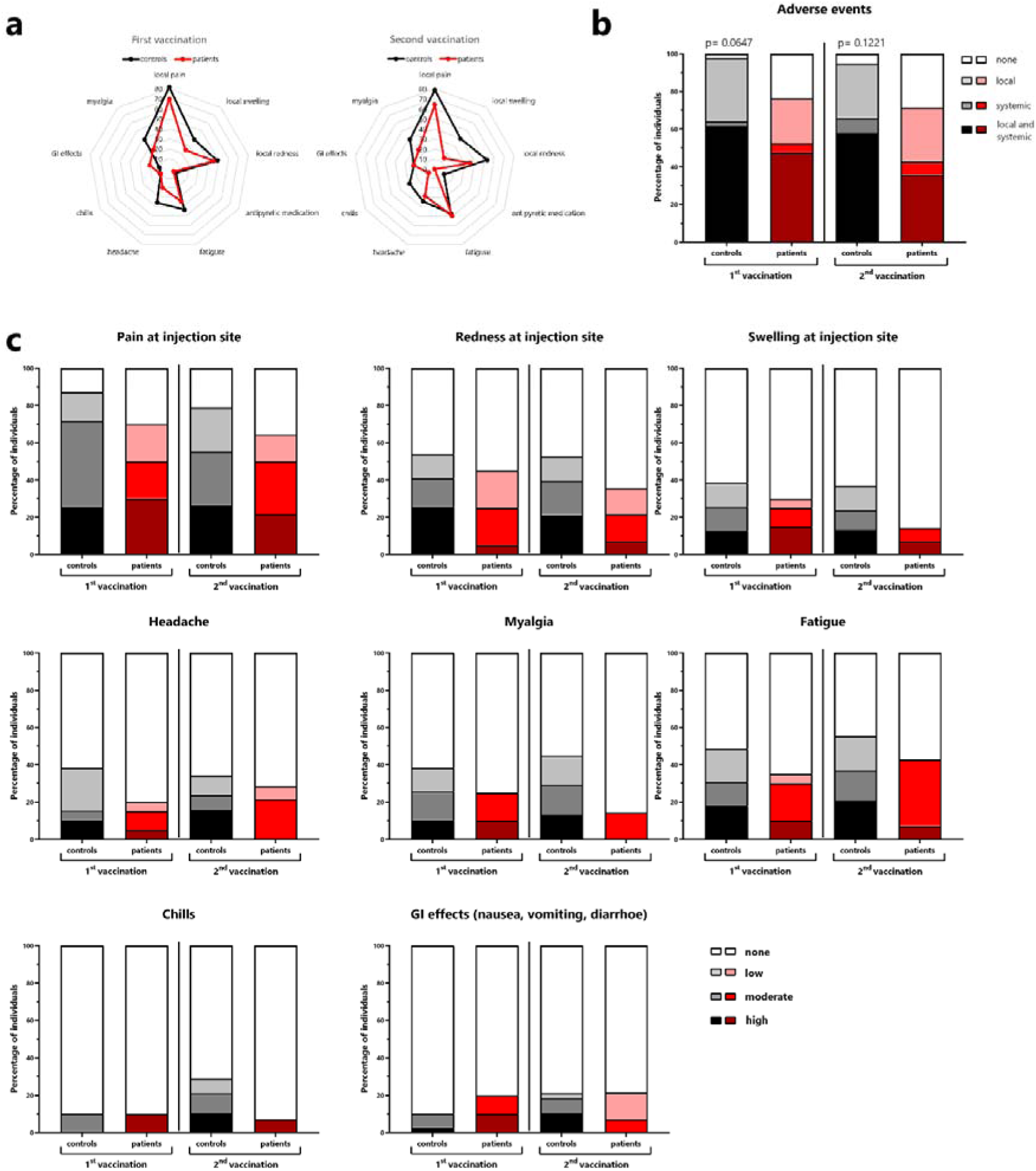
Low reactogenicity in dialysis patients compared to controls. Reactogenicity within the first week after each vaccine dose was self-reported using a standardized questionnaire. **(a)** The distribution of local and systemic adverse events in healthy controls and dialysis patients after the first and second vaccination. **(b)** Percentage of subjects who experienced no adverse events, only local adverse events, only systemic adverse events, or both. **(c)** Adverse events perceived by individuals were ranked and classified by severity (none, low, moderate, high). Comparisons between groups were analyzed using the X^2^ test.

## Discussion

The recombinant vaccine HZ/su was approved for the prevention of herpes zoster and associated serious complications in immunocompromised patients such as dialysis patients, who are at increased risk of VZV reactivation due to uremic immunodeficiency^9, 10^. Knowledge on the immunogenicity and reactogenicity in dialysis patients in relation to immunocompetent controls was limited. In this observational study, we show that two standard doses of the HZ/su vaccine were well tolerated and induced a multifunctional VZV-specific CD4 T-cell response in dialysis patients. In contrast, CD8 T-cells were only poorly induced. At the humoral level, we also observed an increase in VZV-specific IgG antibodies and neutralizing activity. Overall, both cellular and humoral immune responses were slightly less pronounced in patients than in immunocompetent controls, and long-term levels of VZV-specific CD4 T-cells and IgG after one year were less durable.

Our results are consistent with previous observations after VZV-vaccination of healthy controls^4, 22, 30, 31^. We now show that the HZ/su vaccine induced a significant increase in VZV-specific CD4 T-cell levels in dialysis patients. We have previously shown that VZV reactivation in patients with herpes zoster or meningitis leads to an upregulation of CTLA-4 on VZV-specific T-cells, which normalized after resolution of symptoms^27, 28^. Similar dynamics of CTLA-4 expression was now also observed during vaccine-induced T-cells and thereafter. This suggests that CTLA-4 expression on VZV-specific CD4 T-cells may represent a direct measure for recent antigen encounter resulting from either infection or vaccination. In contrast, the cytokine profile seems to differ during infection and vaccination, as vaccine-induced VZV-specific CD4 T-cells showed a multifunctional Th1 phenotype, whereas Th1 CD4 T-cells after acute zoster were functionally restricted and predominantly expressed IFNγ only^28^. In general, the quantitative and functional increase in vaccine-specific CD4 T-cells after the second vaccination may contribute to better protection through proliferation and secretion of effector molecules^32^ and emphasizes the importance of the booster dose to establish a robust immune response. However, compared to controls, dialysis patients mounted significantly lower VZV-specific CD4 T-cell levels after both vaccinations which remained lower at one year follow-up. Phenotypical analysis of vaccine-induced CD4 memory cells revealed a higher proportion of central memory T-cells and lower proportion of effector memory T-cells in patients compared to controls. Similar observations of an impaired effector memory CD4 T-cell response were made in dialysis patients two weeks after hepatitis B vaccination^33^. This may result from the fact that dialysis patients have a higher apoptosis rate of T-cells^34, 35^. As central memory T-cells are better protected against apoptosis^36, 37^, apoptosis may preferentially affect effector memory T-cells in patients.

Compared to the VZV-specific CD4 T-cell levels, specific CD8 T-cells were poorly induced in both dialysis patients and controls, and were primarily detected after proliferation upon longer stimulation times. This is consistent with other studies on the HZ/su vaccine^31, 38^, and is a typical feature of protein-based vaccines in general, which are predominantly presented via the MHC class II pathway and thus primarily activate CD4 T-cells. This is also illustrated by the protein-based SARS-CoV-2 vaccine NVX-CoV2373, where vaccine-induced CD8 T-cell levels were significantly lower as compared to vector-based or mRNA vaccines^25, 39, 40^. Based on the observation that the HZ/su vaccine is highly effective^21, 22^, CD4 T-cells may have a dominant role in mediating protection from herpes zoster^41^. Interestingly, despite low levels of VZV-specific CD8 T-cells after short-term stimulation ex vivo, proliferation of specific CD8 T-cells became detectable after 7 days, albeit to a lesser extent than CD4 T-cells. It is possible that protein-derived peptides are presented to MHC class I via cross-presentation and recognized by CD8 T-cells. The AS01_B_ adjuvant used for HZ/su contains MLP/QS21, which is known to improve antigen cross-presentation^42^ may thereby induce CD8 T-cells and also increase their cytotoxic activity^43^.

Although the induction of the humoral VZV response appears to play a less important role in preventing VZV reactivation^44, 45^, it is crucial in preventing reinfection with varicella^5, 46^. In line with a high VZV-seroprevalence in the world population^47^, the majority of individuals in our study were already seropositive before the first vaccination. In both patients and controls, the HZ/su vaccine led to a further induction of VZV-specific IgG with strong neutralization activity. However, as with CD4 T-cells, the increase in specific IgG were less pronounced in dialysis patients and were less stable over time. In our study, we characterized the circulating fraction of Tfh-cells in the blood as surrogate population of Tfh-cells that interact with B-cells in secondary lymphoid organs and provide support in the production of high-affinity antibodies. However, we were unable to reveal pronounced vaccination-induced changes, which may be due to the fact that Tfh-cell levels were identified in an antigen-non-specific manner, and that results from circulating Tfh-cells may not reflect dynamic changes in the lymph nodes. Likewise, no vaccine-induced changes in plasmablasts of dialysis patients and controls were detectable in circulation. However, dialysis patients generally showed significantly lower numbers of B-cells with an imbalance in B-cell subpopulations such as higher levels of naive B-cells and lower levels of plasmablasts and switched memory B-cells, which may contribute to the state of uremic immunodeficiency^48, 49^. Similar differences in the distribution of B-cell subpopulations have also been described in other diseases such as systemic sclerosis^50^. One reason for the higher percentage of naive B-cells may be increased production to maintain homeostasis, while a lower percentage of non-switched B-cells in patients is possibly caused by increased apoptosis^51^.

In our study, we show that the HZ/su vaccine was well tolerated by dialysis patients, which is in line with a recent adherence and safety study^52^. Local adverse events were more frequently reported as compared to systemic reactions, and overall less frequent than in controls, which has already been observed with vaccinations against SARS-CoV-2^53^. This may be explained by the fact that dialysis patients often suffer from chronic complaints such as fatigue or headaches due to their uremic disease and the dialysis procedure^54^, so that vaccination reactions, especially systemic ones, are perceived as less severe. Thus, both the good immunogenicity as well as the low reactogenicity of the vaccine may increase compliance to adherence with vaccine recommendations and thus contribute to a reduction in cases of herpes zoster and postherpetic neuralgia.

The strength of our study is the investigation of both humoral and cellular immunity including phenotypical characterization of VZV-specific CD4 T-cells in direct comparison with immunocompetent controls, which to our knowledge has not been investigated before. Similar results have been reported in transplant recipients^24^, although no control group was included to specifically analyze the effect of immunodeficiency. Limitations include the low overall number of individuals and lack of follow-up beyond one year, which does not provide information on effectiveness. Nevertheless, sample size was sufficient to characterize immunogenicity and reactogenicity, and reveal differences between controls and patients. Unlike after SARS-CoV-2 vaccination, HZ/su induced a specific CD4 T-cell response and IgG in the majority of patients after two doses of vaccine^55, 56^. It is very likely that dialysis patients benefit from the previous contact with VZV and thus from a pre-existing immune response that was boosted by the two HZ/su vaccine doses. In a study with healthy individuals, robust immunogenicity of HZ/su has already been shown to persist over a period of 10 years^57^. Similar long-term follow-up data are not yet available for immunocompromised individuals but are important in light of our findings that cellular and humoral immunity appears less stable in dialysis patients than in controls. A more rapid loss of vaccine-induced immune responses has already been described after influenza vaccination of dialysis patients, while the immune response in healthy individuals remained more stable^15^. While yearly influenza vaccination is recommended, future studies should clarify whether dialysis patients may benefit from additional doses of HZ/su vaccines. On an individual level, future studies should determine thresholds for cellular and/or humoral immune response parameters associated with increasing risk of VZV reactivation.

## Data Availability

All data produced in the present study are available upon reasonable request to the authors

## Data sharing statement

All figures and tables have associated raw data. The data that support the findings of this study are available from the corresponding author upon request.

## Author Contributions

F.H., T.S., D.S., M.E., U.S., and M.S. designed the study and the experiments, F.H. and D.S performed experiments; F.H., S.L., M.G., K. B., J.M., U.S., and M.S. contributed to study design, patient recruitment, and clinical data acquisition. F.H. and M.S. performed statistical analysis. T.S., D.S., and M.S. supervised all parts of the study; F.H., and M.S. wrote the manuscript. All authors approved the final version of the manuscript.

## Supplementary material

Hielscher_KI_supplement.pdf

### Supplementary methods

**Supplementary table S1:** Underlying diseases in dialysis patients

**Supplementary Figure S1:** Schematic representation of study design. Patients and controls received two doses of the Hz/su vaccine, and blood samples were drawn before each vaccination as well as two weeks after the first and the second vaccination. In addition, a final blood sample was drawn 12 months after the first vaccination (i.e. 9 months after the second). Adverse events were self-reported within the first week after the first and the second vaccination using a standardized questionnaire.

**Supplementary Figure S2**: Cytokine expression of VZV-specific and SEB-reactive CD4 T-cells. **(a)** VZV-specific or **(b)** SEB-reactive CD4 T-cells were stimulated and CD69-positive CD4 T-cells producing IL-2, TNF or IFNγ were quantified after the first, the second vaccination and on follow-up one year after the first vaccination. VZV-specific CD4 T-cell levels are displayed after subtraction of negative control values. Samples from healthy controls (red) and dialysis patients (grey) were compared at each time point. Bars represent median values with interquartile ranges. Statistical analysis was performed using Mann-Whitney test. IL-2, interleukin 2; IFN, interferon; TNF, tumor necrosis factor, VZV, *Varicella zoster virus*; SEB, *Staphylococcus aureus* enterotoxin B.

**Supplementary Figure S3:** CD4 and CD8 T-cell differentiation in dialysis patients and controls. Differentiation status of bulk **(a)** CD4 T-cells or **(b)** CD8 T-cells classified into naive, central memory (CM), effector memory (EM), and terminally differentiated effector memory (TEMRA) cells based on expression of CD45RO and CD27. T-cell populations were compared between controls (gray) and dialysis patients (red) after the first and the second vaccinations and on follow-up. Statistical analysis was performed using Mann-Whitney test. Bars represent medians with interquartile ranges.

**Supplementary Figure S4:** Characterization of follicular T helper cells. **(a)** Representative plots of follicular T helper (Tfh) cells identified as CXCR5+ CD4 T-cells. Tfh-cells were analyzed for expression of PD-1 and ICOS, and the differentiation into naive, central memory (CM), effector memory (EM), and terminally differentiated effector memory (TEMRA) was determined using CD45RO and CD27. **(b)** The percentage of Tfh-cells among CD4 T-cells, and the median fluorescence intensity (MFI) of **(c)** ICOS and **(d)** PD-1 were determined over time in controls (black) and dialysis patients (red). Bold lines represent medians and Friedman test was performed for paired analyses. **(e)** The differentiation status of Tfh-cells before the first vaccination, after the second vaccination and after one year (follow-up) is shown in controls (grey) and patients (red). Bars represent medians with interquartile ranges. ICOS, inducible T-cell costimulator; PD-1, Programmed cell death protein 1.

**Supplementary references**

## Acknowledgements

The authors thank Candida Guckelmus, Rebecca Urschel and Silvia Meier for their excellent technical support. We would also like to thank all participants in this study. Financial support was provided in part by HOMFORexzellent (to D.S.).

