## Supplementary_information for "The inactivated herpes zoster vaccine HZ/su induces a varicella zoster virus specific cellular and humoral immune response in dialysis patients"

Running head: VZV-specific immunity in dialysis patients

Franziska Hielscher, MSc, Tina Schmidt, PhD, Martin Enders, MD, Sarah Leyking, MD, Markus Gerhart, MD, Kai van Bentum, MD, Janine Mihm, MD, David Schub, PhD, Urban Sester, MD, and Martina Sester, PhD

This supplement contains supplementary methods, a supplementary table S1 and 4 supplementary figures S1-S4.

### Supplementary methods

#### Quantification of lymphocyte subpopulations

To quantitate lymphocyte subpopulations 100µl heparinized whole blood was washed once with RPMI, stained for characteristic phenotypic markers and analyzed using flow cytometry as described before^1^. To identify follicular helper T-cells, antibodies against CD4 (clone SK3, 1:5.5) and CXRC5 (clone RF8B2, 1:5.5) were used. In addition, T-cells were analyzed for the surface markers ICOS (clone DX29, 1:3.65) and PD-1 (clone MIH4, 1:3.65). B-cells were identified as CD19^+^ (clone HIB19, 1:16.7) CD3^-^ (clone SK7, 1:26.7) and their differentiation status was determined using antibodies against IgD (clone IA6-2, 1:20) and CD27 (clone L128, 1:3.3). Among switched-memory B-cells, plasmablasts were identified as CD38^+^ (clone HB-7,1:33.3).

#### Quantification and characterization of VZV-specific T-cells

The analysis of antigen-specific T-cells was carried out as previously described for other VZV antigens^2, 3^. In brief, 450µl heparinized whole blood was stimulated with 2µg/ml overlapping VZV gE peptides (Swiss-Prot ID: P09259, JPT, Berlin, Germany). As a positive control, blood samples were stimulated with 2.5 μg/ml *Staphylococcus aureus* enterotoxin B (SEB) (Sigma-Aldrich, St. Louis, MO, USA), while 0.56% DMSO served as a negative control. All stimulations were performed in the presence of 1 μg/ml anti-CD28 and anti-CD49d antibodies (BD Biosciences, San Jose, CA, USA). After 2 hours of incubation, 10 µg/ml brefeldin A was added for intracellular cytokine accumulation. After an additional 4 hours, cells were fixed and immunostaining was performed using anti-CD4 (clone SK3, 1:33.3), anti-CD8 (clone SK1, 1:12.5), anti-CD69 (clone L78, 1:33.3), anti-IFNγ (clone 4S.B3, 1:100), anti-IL-2 (clone MQ1-17H12, 1:16.7), anti-TNF (clone MAb11, 1:20), and anti-CTLA-4 (clone BNI3, 1:50). To characterize VZV-specific memory T-cells, immunostaining was performed using anti-CD4 (clone SK3, 1:100), anti-CD69 (clone L78, 1:25), anti-IFNγ (clone 4S.B3, 1:100), anti-CD45RO (clone UCHL-1, 1:40), and anti-CD27 (clone M-T271, 1:20). Flow cytometric analyses were performed on FACS Canto II, using FACSDiva Software 6.1.3 (BD). VZV-specific CD4 or CD8 T-cells were identified as activated CD69-positive T-cells producing IFNγ and further characterized for expression of cell surface markers and additional cytokines. Percentage of VZV-specific CD4 and CD8 T-cells were determined by subtracting the corresponding negative controls.

#### Proliferation activity of VZV-specific T-cells

VZV-specific proliferation was analyzed using carboxyfluoresceindiacetate-succinimidylester (CFDA-SE) assay, as previously described^4^. In brief, peripheral blood mononuclear cells (PBMC) were isolated by Ficoll density gradient (Linaris) and stained with CFDA-SE (5µM, Invitrogen). Cells were cultured at 2x10^7^ cells/ml in RPMI-5% FCS-1% antibiotics in the presence of 2µg/ml overlapping VZV gE peptides (Swiss-Prot ID: P09259, JPT, Berlin, Germany). Negative or positive control stimulations were carried out using 0.56% DMSO or 2.5 μg/ml SEB, respectively. Cells were incubated at 37°C and 5% CO_2_. SEB stimulated cells were splitted (1:1) on day 3 or 4. Flow cytometric analysis was performed after 7 days of proliferation after co-staining using antibodies towards CD3 (clone SK7, 1:50), CD4 (clone SK3, 1:12.5), CD8 (clone SK1, 1: 8.33).

#### Analysis of VZV-specific IgG antibodies and neutralization activity

VZV-specific antibodies were quantified using a commercial anti-IgG enzyme-linked immunosorbent assay (Euroimmun AG, Lübeck, Germany). IgG levels < 80 IU/L were scored negative, levels 80–110 IU/L were scored intermediate, and levels > 110 IU/L were scored positive according to the manufacturer’s instructions.

To analyze the functionality of VZV-specific antibodies, a neutralization test was carried out. The assay was performed in duplicate using serial dilutions of serum samples in E-MEM-2% FCS-0.1% antibiotics. Subsequently, 100µl VZV (Clade 3, strain Nr. 13, Original-Nr: 1219/07; kindly provided by Prof. Dr. med. Hartmut Hengel; reference laboratory for HSV/VZV; Freiburg; Germany) diluted 1:40 in medium was added and the samples were incubated for 90 minutes at 37°C and 5% CO_2_. Finally, 100µl embryonic lung fibroblasts (100,000 cells/ml) were added and plates were incubated for 5 days. Serial dilutions without serum served as positive control and a dilution series with cell suspension only served as negative control. The cells were fixed for 10 minutes with 100µl ice-cooled acetone/methanol (40:60). Anti-VZV antibody towards the immediate early gene 62 (MAB 8616, 1:1000, Sigma-Aldrich) was added as primary antibody. Goat anti-Mouse IgG (1:200, Thermo Scientific) was used as secondary antibody. After adding 50µl AEC substrate (Sigma-Aldrich) and incubating for 30 minutes at 37°C and 5% CO_2_, the reaction was stopped with distilled water. Finally, plaques were counted microscopically, and the geometrical mean (GM) was determined.

### Supplementary table

#### Table S1: Underlying diseases in dialysis patients

| **Underlying disease** | **n (%)** |
| --- | --- |
| Chronic glomerulonephritis | 4 (13.8) |
| Hereditary nephropathy | 4 (13.8)^$^ |
| Urological nephropathies (obstructive, nephrectomy) | 4 (13.8) |
| Diabetic nephropathy | 6 (20.7) |
| Hypertensive/vascular nephropathy | 6 (20.7) |
| Others | 5 (17.2)^#^ |

^$^incl. autosomal dominant polycystic kidney disease; ^#^incl. lipidapheresis (n=1).

### Supplementary figures

#### Figure S1

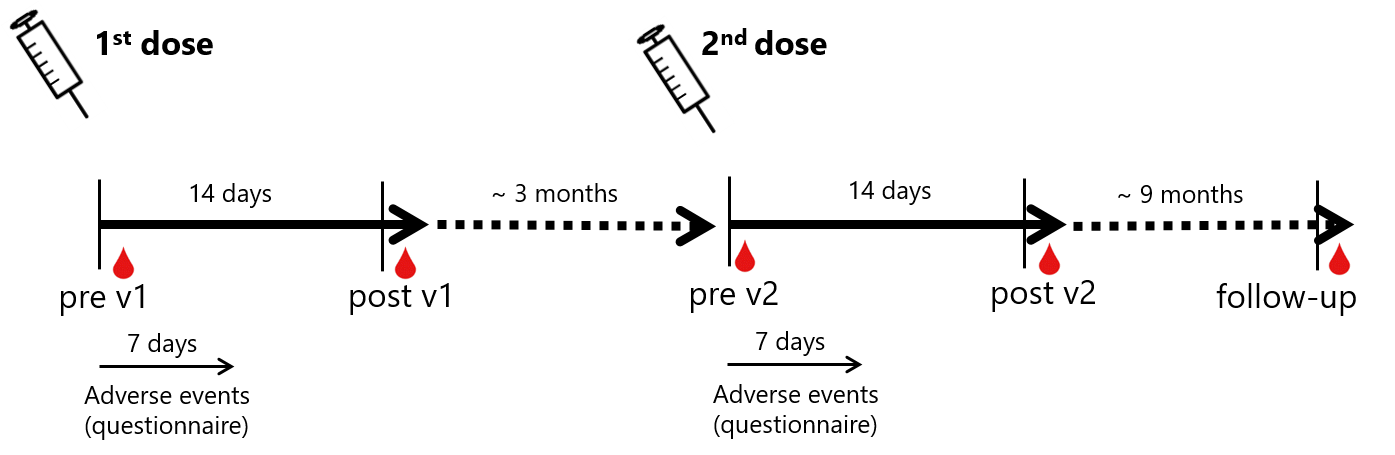

**Figure S1: Schematic representation of study design.** Patients and controls received two doses of the Hz/su vaccine, and blood samples were drawn before each vaccination as well as two weeks after the first and the second vaccination. In addition, a final blood sample was drawn 12 months after the first vaccination (i.e. 9 months after the second). Adverse events were self-reported within the first week after the first and the second vaccination using a standardized questionnaire.

#### Figure S2

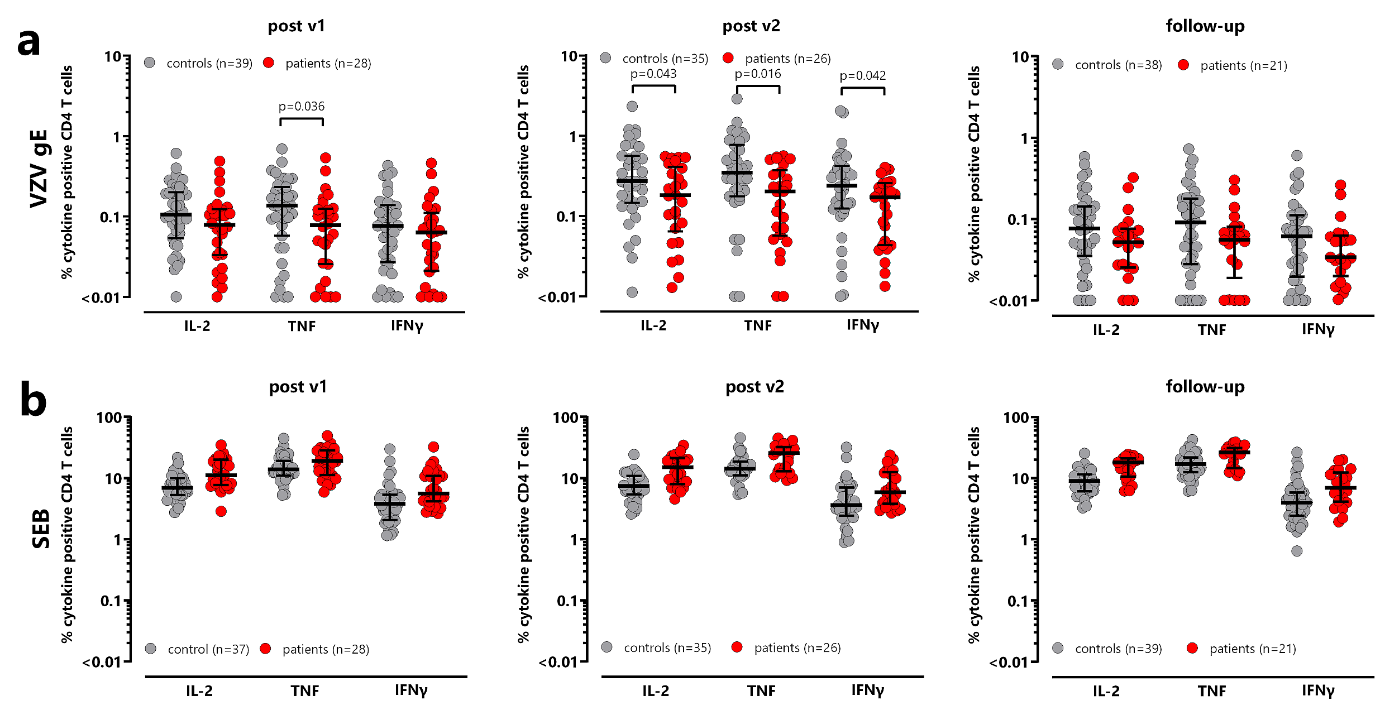

**Figure S2: Cytokine expression of VZV-specific and SEB-reactive CD4 T-cells. (a)** VZV-specific or **(b)** SEB-reactive CD4 T-cells were stimulated and CD69-positive CD4 T-cells producing IL-2, TNF or IFNγ were quantified after the first, the second vaccination and on follow-up one year after the first vaccination. VZV-specific CD4 T-cell levels are displayed after subtraction of negative control values. Samples from healthy controls (red) and dialysis patients (grey) were compared at each time point. Bars represent median values with interquartile ranges. Statistical analysis was performed using Mann-Whitney test. IL-2, interleukin 2; IFN, interferon; TNF, tumor necrosis factor, VZV, *Varicella zoster virus*; SEB, *Staphylococcus aureus* enterotoxin B.

#### Figure S3

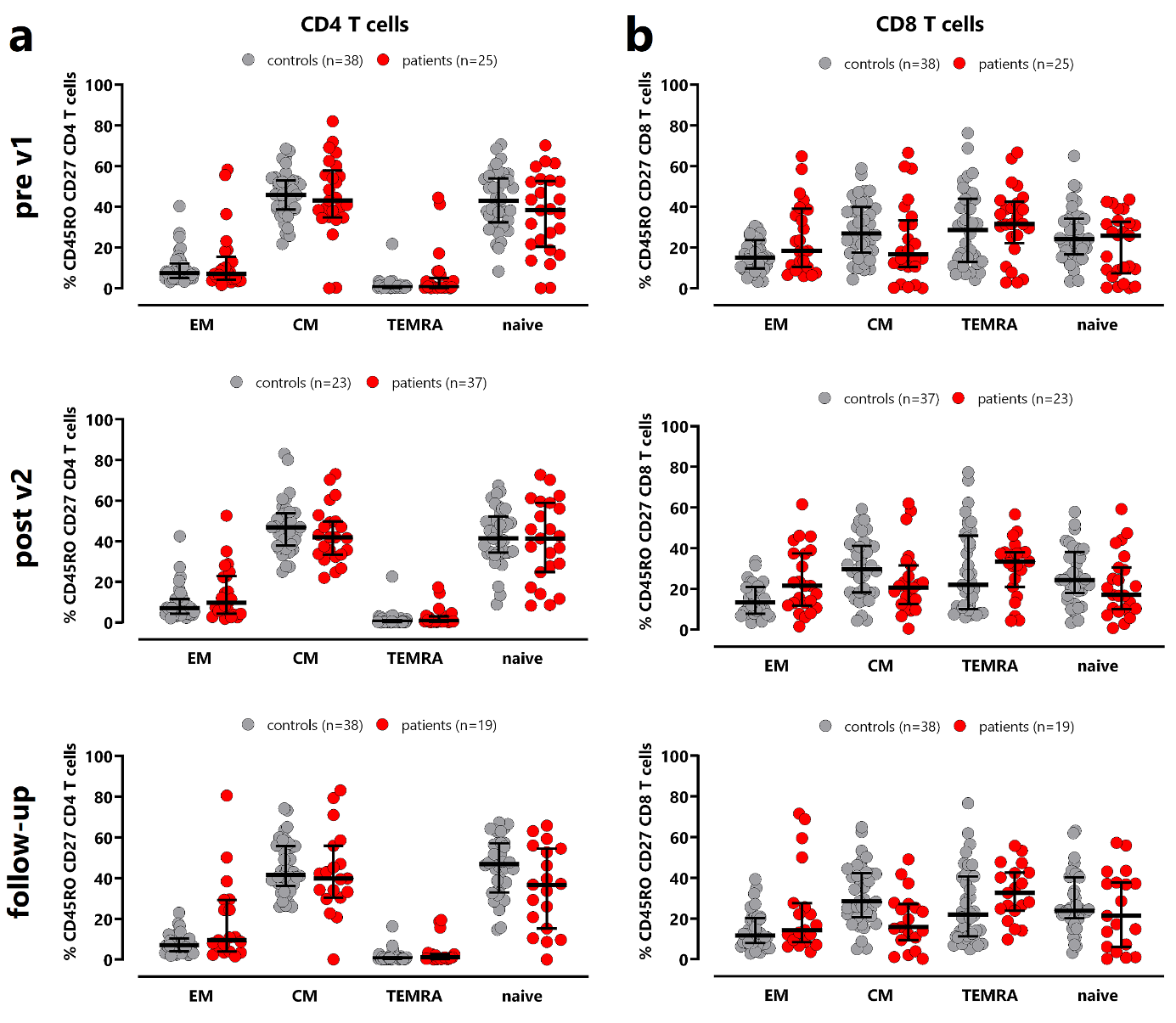

**Figure S3: CD4 and CD8 T-cell differentiation in dialysis patients and controls.** Differentiation status of bulk **(a)** CD4 T-cells or **(b)** CD8 T-cells classified into naive, central memory (CM), effector memory (EM), and terminally differentiated effector memory (TEMRA) cells based on expression of CD45RO and CD27. T-cell populations were compared between controls (gray) and dialysis patients (red) after the first and the second vaccinations and on follow-up. Statistical analysis was performed using Mann-Whitney test. Bars represent medians with interquartile ranges.

#### Figure S4

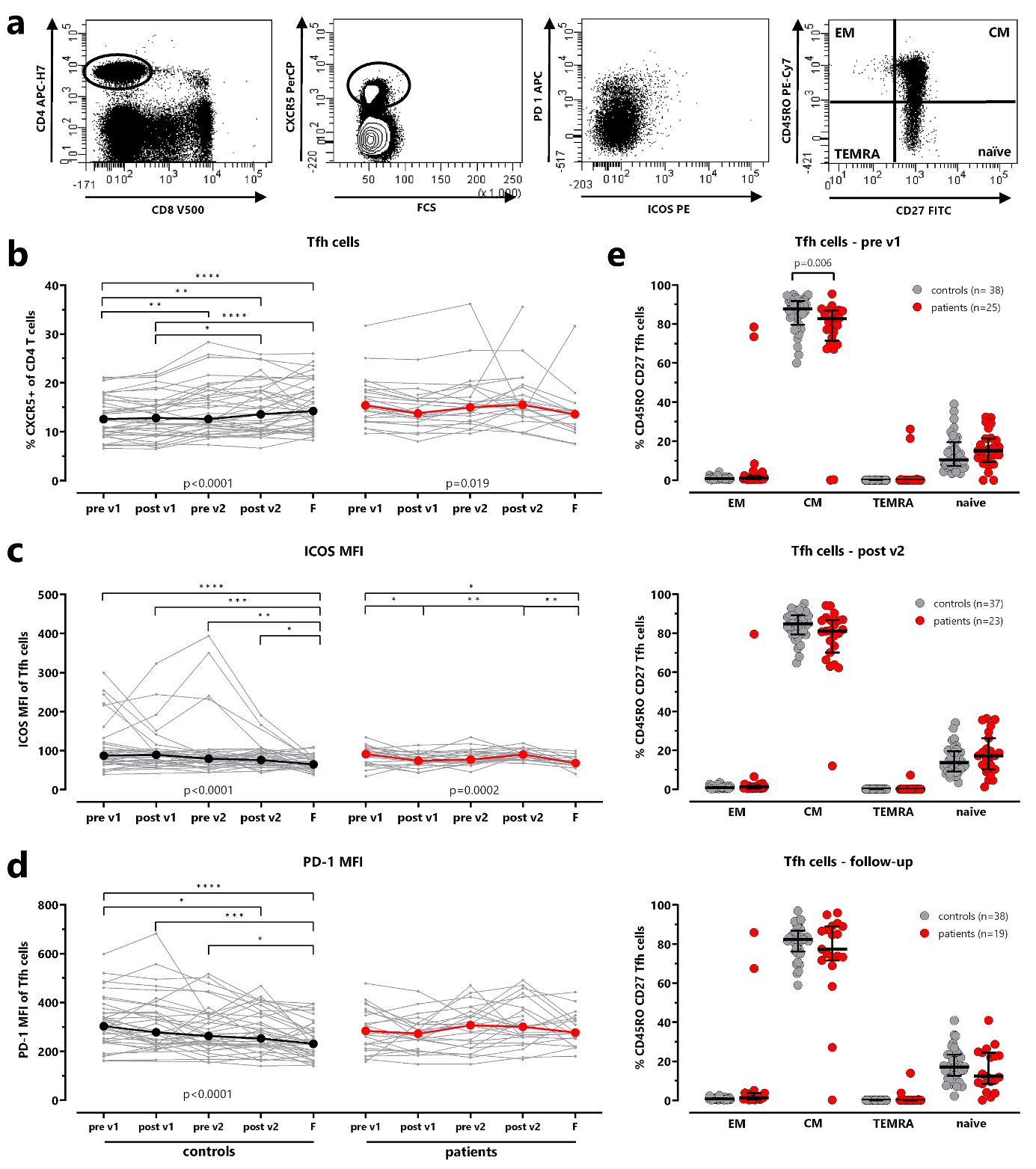

**Figure S4: Characterization of follicular T helper cells. (a)** Representative plots of follicular T helper (Tfh) cells identified as CXCR5+ CD4 T-cells. Tfh-cells were analyzed for expression of PD-1 and ICOS, and the differentiation into naive, central memory (CM), effector memory (EM), and terminally differentiated effector memory (TEMRA) was determined using CD45RO and CD27. **(b)** The percentage of Tfh-cells among CD4 T-cells, and the median fluorescence intensity (MFI) of **(c)** ICOS and **(d)** PD-1 were determined over time in controls (black) and dialysis patients (red). Bold lines represent medians and Friedman test was performed for paired analyses. **(e)** The differentiation status of Tfh-cells before the first vaccination, after the second vaccination and after one year (follow-up) is shown in controls (grey) and patients (red). Bars represent medians with interquartile ranges. ICOS, inducible T-cell costimulator; PD-1, Programmed cell death protein 1.
